# Artificial-Intelligence Powered Identification of High-Risk Breast Cancer Subgroups Using Mammography: A Multicenter Study Integrating Automated Brightest Density Measures with Deep Learning Metrics

**DOI:** 10.1101/2024.01.28.24301639

**Authors:** Yeojin Jeong, Jeesoo Lee, Young-jin Lee, Jiyun Hwang, Sae Byul Lee, Tae-Kyung Yoo, Myeong-Seong Kim, Jae Il Kim, John L Hopper, Tuong L Nguyen, Jong Won Lee, Joohon Sung

**Affiliations:** Genome & Health Data Lab, Seoul National University School of Public Health, Seoul, Korea; Department of Surgery, Asan Medical Center, University of Ulsan College of Medicine, Seoul, Republic of Korea; Department of Radiology, National Cancer Center, Goyang-si, Korea; Department of Surgery, Inje University Ilsan Paik Hospital, Goyang-si, Korea; Department of Biostatistics and Epidemiology, School of Global and Population Health, University of Melbourne, Melbourne, Australia

**Keywords:** Breast cancer risk, Deep learning, Digital mammography, Multi-level mammographic breast density, High-risk group identification

## Abstract

Mammography plays a crucial role in breast cancer (BC) risk assessment. Recent breakthroughs show that deep learning (DL) in mammography is expanding from diagnosis to effective risk prediction. Moreover, the brightest mammographic breast density (MBD), termed “cirrocumulus,” signifies an authentic risk. Addressing the challenges in quantifying above recent measures, we present MIDAS: a DL-derived system for multi-level MBD and risk feature score (FS). Using >260,000 multicenter images from South Korea and the US, FS consistently outperforms conventional MBD metrics in risk stratification. Only within the high FS, cirrocumulus further enriches assessment, pinpointing “double-higher” subgroup. Their risk profiles are notable: women in double upper-tertile showed OR=10.20 for Koreans and 5.67 for US, and OR=7.09 for scree-detected cases (US only). We also reveals the “black-box” nature of FS that it predominantly captures complex patterns of higher-intensity MBD. Our research enhances the potential of digital mammography in identifying individuals at elevated BC risks.

## 1. Introduction

Breast cancer (BC) stands as the primary cause of women’s malignant neoplasms (Kang et al., 2023; Rajappa et al., 2023). In regions like South Korea and East Asia, a greater proportion of BC cases manifest before the age of 45 compared to European Ancestry populations (National Central Cancer Registry, 2023), underscoring the need for early screening programs targeted at women below 45. While the American Cancer Society (ACS) recommends MRI screening for high-risk individuals (Saslow et al., 2007; Smith et al., 2019), the identification of such high-risk groups remains a challenge due to the moderate predictive power of existing risk assessment tools (Terry et al., 2019). Recent endeavors to improve these risk assessment models have integrated information from mammography and polygenic risk scores (Lee et al., 2019). Mammographic Breast Density (MBD) is a well-established risk factor of BC (Boyd et al., 2007; Brentnall et al., 2015; Britt et al., 2020); however, MBD is not as commonly incorporated into risk prediction models as clinical and epidemiological risk factors. To effectively leverage MBD data, two key elements are needed: mammographic images and methods to accurately measure MBD. Conventionally, MBD has been gauged manually by experts, often aided by software like Cumulus (Byng et al., 1994). Although automated measurement techniques exist for diverse mammography modalities and vendors (Alonzo-Proulx et al., 2010; Haji Maghsoudi et al., 2021; Keller et al., 2015), their deployment and widespread use in clinical settings have been limited due to compatibility or copyright issues.

While the elevated risk of BC in women with higher MBD levels is well-established, there’s ongoing debate about which MBD metrics most accurately signify this risk. For instance, traditional MBD levels show a markedly reduced correlation with screen-detected BC but not with interval cases, hinting at a “masking effect” (Mandelson et al., 2000). With the broader adoption of full-field digital mammography (FFDM), new indices like “altocumulus” and “cirrocumulus”, representing areas brighter than white, have been proposed (Nguyen et al., 2017; Nguyen et al., 2018b). For clarity, we’ll use these meteorological-inspired terms (cumulus, altocumulus, and cirrocumulus) to signify MBD levels that discriminates conventional MBD measures by intensity. Notably, “cirrocumulus” (the brightest area) has demonstrated a significant association with screening-detected cancers (Nguyen et al., 2018a; Wanders et al., 2017), implying that traditional MBD, or “cumulus,” may actually serve as a confounder, obscuring the genuine risk signal from cirrocumulus. As of now, however, multilevel measurements are conducted exclusively through expert manual evaluation.

Deep learning (DL) applications in mammography initially focused on detecting cancerous lesions (Lotter et al., 2021). Subsequently, DL techniques have been honed and validated to exceed the performance of traditional breast cancer (BC) risk prediction models, using mammography images of healthy women (Yala et al., 2022; Yala et al., 2021). Numerous studies corroborate that risk prediction models enriched with DL-derived features outperform traditional models or those that include mammographic breast density (MBD) (Arefan et al., 2020; Dembrower et al., 2020; Yala et al., 2019; Zhu et al., 2021). Despite these advancements, the specific mammographic characteristics or features that DL identifies as high-risk remain poorly understood, limiting its further applications. To understand the relevance of specific features, attempts such as “explainable AI (XAI)”, often described as a “unboxing black box”, are being made to demystify DL’s inner workings, including efforts to understand the meaning of specific features (Kim, 2017). Our study aims to unravel the specific DL features associated with elevated breast cancer risk by applying XAI and comparing those with multi-level MBD; we attempt to provide insights into the meaning to these otherwise opaque algorithms.

Our study introduces MIDAS (Mammography-based Integrated Density and Risk Analysis System), a platform that estimates DL-derived multi-level MBD measurements and risk feature scores (FS). Utilizing a multi-center mammography database, we developed automated measurements for multi-level MBD and FS using the mammograms. Our ultimate aim is to examine whether we can identify the high-risk individuals using the mammography alone. More specifically, we inspect how the integration of multi-level MBD and FS can amplify the identification of individuals at elevated risk for BC. We rigorously scrutinize and refine each step of our DL methods, from quality control and image processing to *post hoc* analyses aimed at providing further insights into FS.

## 2. Materials and Methods

### 2.1 Analytic flow

Our analytical approach (MIDAS) integrated two autonomous modules: estimation of multi-level pMBD and FS derived from DL. For first module, FFDM images with multi-level MBD annotation by expert were preprocessed, followed by 3-level MBD ground truth mask generation. The image – mask pairs were used to train DL model for semantic segmentation. Multi-level MBDs were measured by using trained DL model, resulting in multi-level pMBD (percent of MBD to total breast area) measurements. For second module, FFDM images with clinical diagnosis from pathological report were preprocessed and then used to train DL model to estimate FS. Finally, FS and multi-level pMBD were jointly utilized to detect a high-risk subgroup of BC. XAI was applied to visually inspect the relevant features of DL model in second module. Fig. 1 provides an overview of the analytical process.

**Fig. 1.**
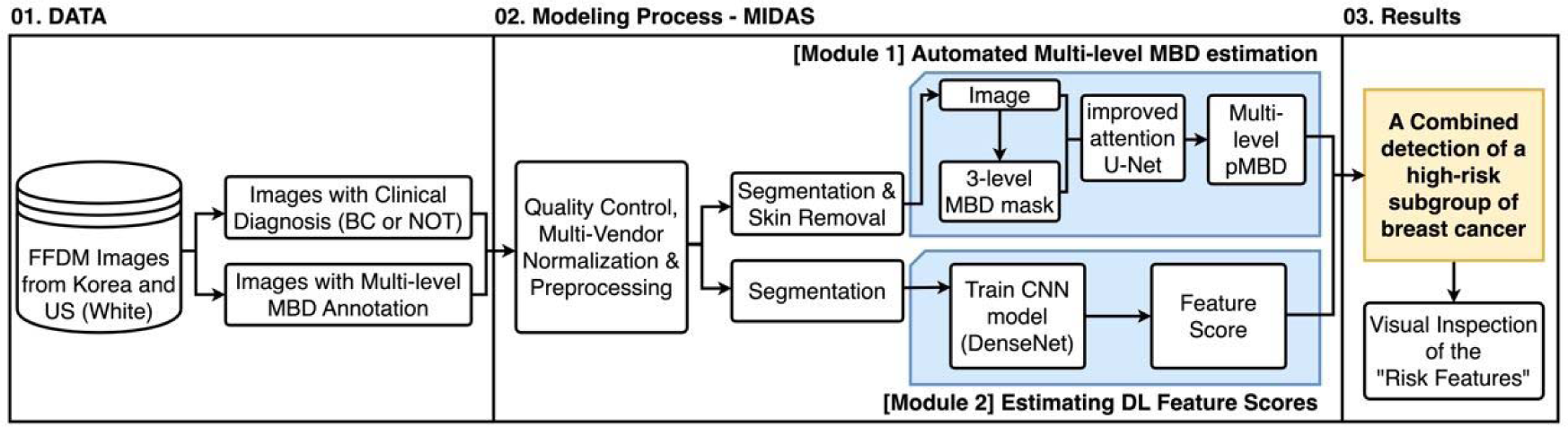
An outline of overall analytic flow. Images with multi-level MBD annotation were used to train DL model for semantic segmentation. Multi-level pMBD were calculated using predicted MBD measurements (module 1). Images with clinical diagnosis from pathological report were used to train DL model to estimate FS (module 2). FS and pMBD were combined to detect a high-risk subgroup of BC.

### 2.2 Data source

FFDM images in raw DICOM format were obtained from five clinical centers in South Korea and database of United States. Five Korean centers include National Cancer Center (NCC), Samsung Seoul Medical Center (SMC), Seoul Asan Medical Center (AMC), Seoul Boramae Medical Center (BMC), and Inje University Ilsan Paik Hospital (IPH), and the Emory Breast Imaging Dataset (EMBED) includes four hospitals mammographic examination data for eight years (Jeong et al., 2023). The images from the Korean Hospitals were acquired during clinical evaluations, thus following a case-control study design; EMBED is a mixture of longitudinal screening data and hospital cases. We selected 2-D FFDM data from all vendors, excluding synthetic or 3-D mammographic images. Both radiological assessments and subsequent pathological diagnoses were annotated for all Korean centers and Emory data, except BMC. For BMC, images classified as BI-RADS category 6 were regarded as BC cases, while those in category 1 were considered controls; all other categories were excluded. Among cases, we excluded images identified as ductal carcinoma in situ (DCIS). We also omitted images identified as benign breast tumors from the training set, regardless of histological types. However, these benign tumors were included in subsequent analyses alongside normal and case images. For consistency, we focused solely on cranio-caudal (CC) views, as expert MBD measurements were exclusively conducted on these. We considered images from the contralateral side of a BC lesion as “case images,” and randomly selected unilateral images from healthy individuals as “control images.” Bilateral breast cancer cases were excluded. For individuals with multiple FFDM images, only baseline images for controls and first-diagnosis images for cases were included in the study. Only two clinical centers’ images had three levels of MBD threshold values measured by experts using previously described methods (Nguyen et al., 2017). These manually assessed multi-level MBD values served as the ground truth for training our automated multi-level MBD measurement system.

### 2.3 Image processing

Overall image processing algorithm operation is depicted in fig 2. The FFDM images first undergo histogram window adjustments to account for vendor-specific differences by normalizing the maximum and minimum pixel values. For this, we utilized Python libraries such as Pydicom (Mason, 2011), NumPy (Harris et al., 2020), and OpenCV (Bradski, 2000). We used a multifactorial dimensionality reduction method of UMAP (McInnes et al., 2020) to show the characteristics of image data between vendors, before and after the normalization. To eliminate markers and other artifacts, we binarized the image and retained only the largest contour, which correspond to the breast area. All the images were reshaped to square by zero-padding the remaining region, then resized to 256 x 256. These resized images were used to train convolutional neural network (CNN) model to estimate “feautre score” (module 2). For images used in automated multi-level MBD measurement (module 1), we applied a method to remove skin from the captured breast region, as the skin is measured as the dense area, thereby confounding MBD estimations. For this, we identified an optimal margin to be removed from training data and a fixed-width margin was eliminated from the periphery of the breast area. Then, we generated 3-level MBD masks by retaining pixels with higher intensity than these experts’ manual measure of multi-level MBD threshold values.

**Fig 2.**
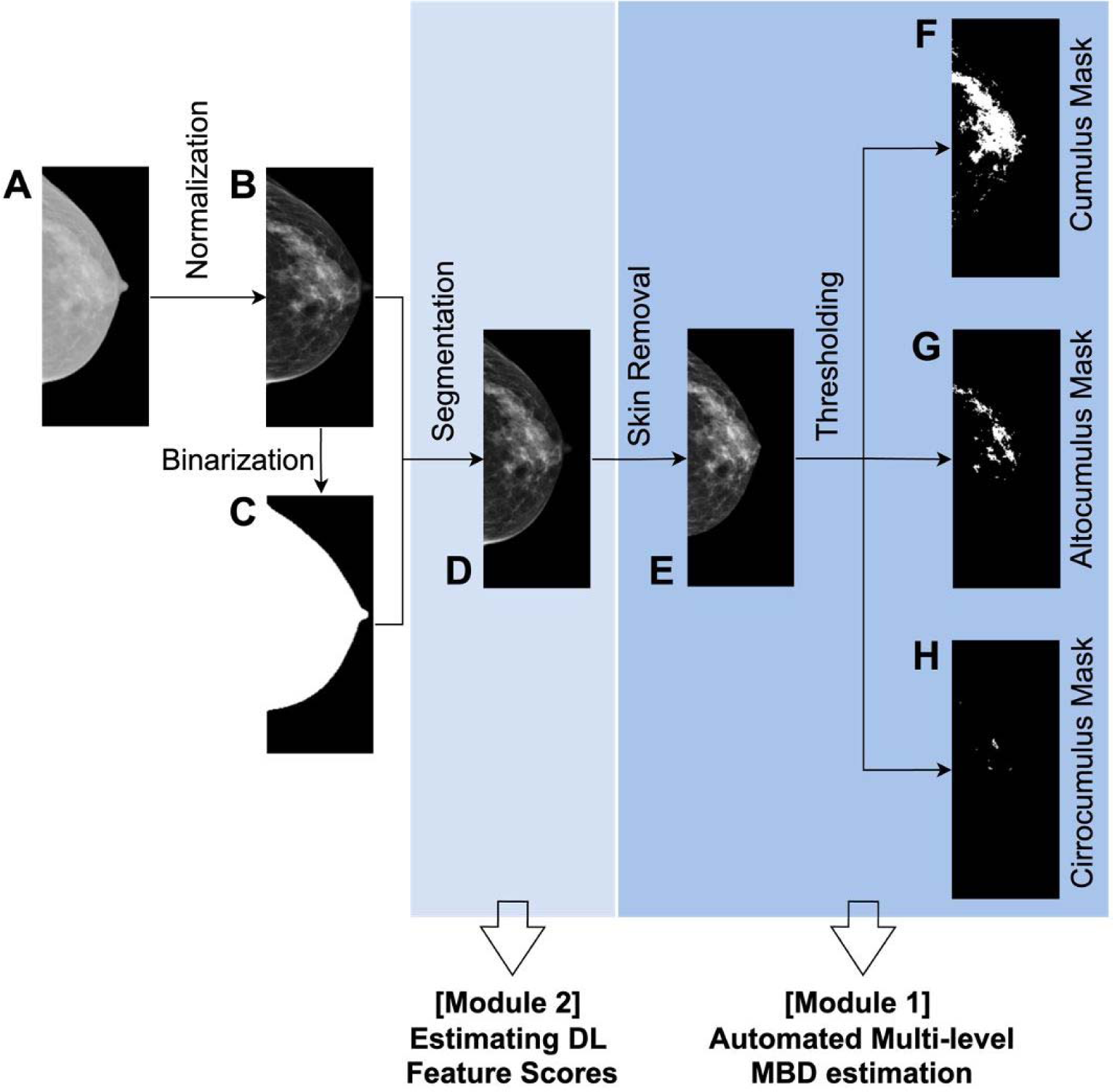
Detailed illustration of FFDM processing for MIDAS. Original FFDM image (panel A) undergo histogram window adjustment, resulting in normalized image (panel B). Largest contour of binarized image (panel C) is overlayed to normalized image for segmentation of breast area (panel D). Images corresponding to panel D are used for training DL model for feature score estimation (module 2). The peripheral margin of breast in image from panel D is removed resulting to the skin removed version of image (panel E). Finally, multi-level MBD masks are generated by only retaining regions with pixels above the threshold measured by expert (panel F: Cumlus mask, panel G: Altocumulus mask, panel H: Cirrocumulus mask). The skin-removed image (panel E) and its corresponding multi-level MBD masks (panel F, G, H) are used for training DL model for automated multi-level MBD estimation (module 1). Note: Only cropped images are shown. Actual processing algorithm includes zero-padding images to square.

### 2.4 MIDAS algorithm operation

#### 2.4.1 Developing automated multi-level MBD measurement

##### Dataset

To create a fully automated method for multi-level MBD measurement, we utilized FFDM images with multi-level MBD (“cumulus”, “altocumulus”, and “cirrocumulus”) annotation evaluated by experts, which consisted of 14,031 FFDM images from two centers in Korea (AMC, NCC). The skin-removed FFDM (Fig 2. E) and its corresponding multi-level MBD masks (Fig 2. (F, G, H)) were used to train CNN for semantic segmentation.

##### Loss function

We used the Tversky Index (TI) to assess segmentation performance. TI serves as a generalized Dice score coefficient (DSC), allowing us to balance between false positive (FP) and false negative (FN) rates, as shown in Equation (1):

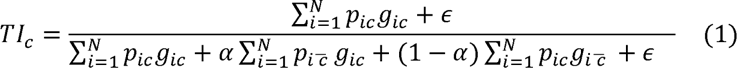

(where, p_ic_ represent the probability that pixel *i* belong of the target class c and non-target class *C*, respectively, and g_ic_ and g_ic_ are similarly defined.)

Hyperparameters α and β can be tuned to shift the emphasis to false positive and false negative detections (In case of α = β = 0.5, TI simplifies to the DSC). The Tversky index is adapted to a loss function (TL) by minimizing ∑_c_ 1- *Ti*c (Hashemi et al., 2018). However, in practice, DL struggles to segment small region of interest (RoI) as they do not contribute to the loss significantly. To alleviate this problem, the authors of improved attention U-Net paper (Abraham and Khan, 2019) proposed the focal Tversky loss function (FTL), which is parameterized by hyperparameter ϒ to control between easy background and hard RoI training examples. The focal parameter exponentiates the cross-entropy loss to focus on hard classes detected with lower probability (Lin et al., 2017). FTL is defined as (equation (2)):

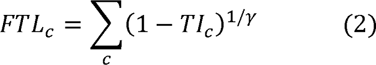

When γ>1, the loss function focuses more on less accurate predictions that have been misclassified.

##### Network architecture

We employed “improved attention U-Net” (Ronneberger et al., 2015b), which is CNN for binary semantic segmentation based on U-Net (Ronneberger et al., 2015a). To mitigate loss of spatial details at the deepest stage of encoding in vanilla U-Net, the attention gates (AG) are added to identify relevant spatial information from low-level feature maps and propagate it to the decoding stage. We used approach of incorporating FTL and TL with improved attention U-Net (Abraham and Khan, 2019) to achieve further balance between precision and recall. In the network, every high dimensional feature in intermediate layers were supervised with FTL (with γ=4/3, recommened by author), with the exception of the last layer which was supervised with TL to avoid over-suppression of loss.

##### Network training

Dataset was randomly splitted into 8: 1: 1 as training: validation: test set, stratifying the vendor. Data augmentation was applied for further improvement: for each training epoch, each image was randomly modified by combinations of elastic deformation; affine transformations including translating, scaling, and rotation; random bright and contrast alteration; and gaussian blur. Since three levels of MBD have substantially different areas for same input image, we trained each segmentation model with different hyperparameter α for TL to find optimal balance between FP and FN. We used α value of 0.1, 0.2,…, 0.9, and choose α for each network which showed maximum DSC between ground truth in test images. All the models were trained with Adam optimizer with initial learning rate 0.0003 and momentum value of 0.9. Details about our deep learning approaches can be found in the supplementary material (Supplementary methods 1).

Using the trained DL model for automated multi-level MBD measurements, we calculated each level of MBD - cumulus, altocumulus, and cirrocumulus. The area of the initially retained largest contour (fig. 2C) was computed as the total breast area. To validate the accuracy of our automated measurements, we compared them to expert evaluations by calculating both Dice score coefficients and correlation coefficients of the measured areas.

#### 2.4.2 Estimating Feature Scores (FS) associated with the risk of breast cancer

To derive “feature scores” (FS) that could discriminate between “case” and “control” images, we utilized a CNN model tailored for binary classification. The dataset (composed of images from fig. 2D) was partitioned into 7:1:2 as training: validation: test set, while maintaining a stratified balance across clinical centers and vendors, as well as preserving the overall case-control ratio. Training was conducted by applying “DenseNet 121” (Huang et al., 2018). To mitigate imbanace between case and control, we applied the case-control ratio as the weight in calculating loss function of binary cross entropy. We conducted model training by 100 epochs and saved the model with highest recall in validation dataset, to prioritize the detection of high-risk individuals. Model was trained with Ranger optimizer (Wright, 2019) (initial learning rate 1e-3). After training the CNN, we calculated the “FS” from the DL model by applying a sigmoid function to the CNN output. Detailed methodology for DL training can be found in Supplementary Methods 2.

### 2.5 Risk prediction of breast cancer by combining multi-level MBD and FS

To assess the individual and combined predictive power of multi-level MBD and FS, we employed several analytical strategies:

1. We first calculated the MBD measures as a “percent of MBD to total breast area” (pMBD) and adjusted them for age and total breast area, following previous studies (Boyd et al., 2006; Checka et al., 2012). We fitted linear regression model, which featured age and total breast area as independent variables, while log-transformed pMBD served as the dependent variable. The residuals were subsequently calculated from fitted linear regression model, and we applied Yeo-Johnson transformation to normalize residuals (Weisberg, 2001). The normalized residuals were then utilized for breast cancer risk prediction (detailed adjustment procedures can be found in Supplementary Method 3).
2. We also calculated MBD adjusted by non-dense area, proxy of BMI adjustment, for comparisons. The adjusted MBD levels were further adjusted by age in the models (Supplementary Method 3).
3. Next, using test dataset (70% for training, 10% for validation, and 20% for test), we determined thresholds for FS and adjusted pMBD based on tertile of control group only. We first evaluated overall trend of BC prevalence along the increase of three levels of adjusted pMBD in groups stratified by FS, using the local regression of moving average. Next, we categorized individuals into nine groups based on established thresholds of both FS and pMBD. We compared the changes in OR according to both pMBD and FS tertiles. We designated individuals in the median tertile of FS as the reference group and calculated odds ratios (OR).
4. The same analysis for tertile groups were conducted according to screen-detected and other cases detected not through regular screening (EMBED only).

### 2.6 *post hoc* analysis of feature scores from DL

We employed “Explainable Artificial Intelligence (XAI)” methods to elucidate the FS produced by our DL models. Utilizing a tailored version of Guided Grad-CAM (Selvaraju et al., 2017), we visualized feature importance of last convolutional layer in DenseNet in relation to both the original mammographic images and multi-level MBD assessments. Our refined Guided Grad-CAM mapping focused on isolating only the more significant features for the comparisons.

## 3. Results

### 3.1 Participants and image data

For Korean data, from an initial pool of 67,749 DICOM images, a refined selection of 13,973 FFDM images met our inclusion criteria. Our study focused on images from major vendors: GE, Hologic, and Lorad. Table 1 describes the demographic and characteristics of the selected data. The average age of the subjects was 52.8 years (s.d. 9.7), and the majority of the images came from GE machines (50.0%), followed by Hologic (40.2%) and Lorad (9.8%). To obtain automated multi-level MBD measurements, we employed images that had been manually annotated for multi-level MBD by experts, regardless of clinical diagnosis. Ground-truth annotations for the three levels of MBD were available for 14,031 cranio-caudal (CC) view images, sourced from AMC and NCC. For EMBED, we used 20% of full EMBED data which is opened. Among the initial 222,880 images, we used 5,022 images of White women were selected. Detailed quality control measures and number of images excluded at each stage are described in fig 3.

**Fig 3.**
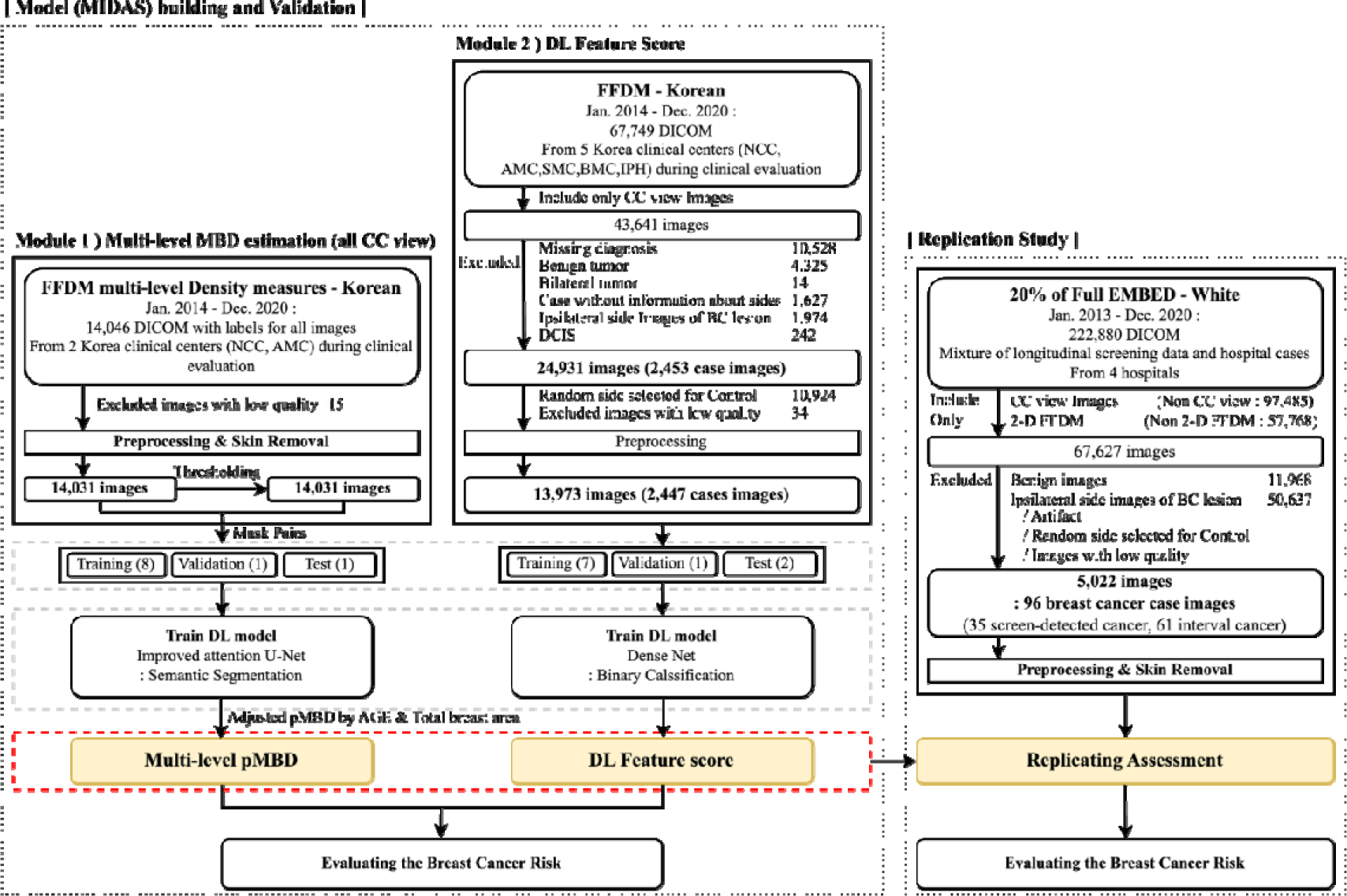
Detailed quality control and analytic process in MIDAS.

**Table 1.** Characteristics of participants and the mammographic images (FFDM) used in this study.

| Clinical<br>Centers | No. of<br>Participants<br>(% case)<br>(Cases /<br>Controls) | No. of<br>Total<br>Images | No of FFDM<br>for Analysis<br>(% case)<br>Case images /<br>Control<br>images | Mean Age<br>(S.D.)<br>Mean age by<br>Case /<br>Control | Ethnicity | Vendors<br>of FFDM | Multi-level<br>MBD Reading<br>by Experts |
| --- | --- | --- | --- | --- | --- | --- | --- |
| Model building and validation |  |  |  |  |  |  |  |
| 1. NCC | 6,752 (21.9%) | 38,541 | 6,760 (21.9%) | 55.2 (9.8) | All | GE (2,537) | 11,956 |
|  | (1,478/ 5,274) |  | (1,478/5,282) | 52.6 (10.0)/ | Korean | Hologic<br>(4,223) |  |
|  |  |  |  | 55.9 (9.5) |  |  |  |
| 2. AMC | 1,849 (26.1%) | 3,785 | 1,849 (26.1%) | 49.6 (5.2) | All | GE (1,849) | 2,075 |
|  | (483/1,366) |  | (483/1,366) | 48.7 (5.0)/ | Korean |  |  |
|  |  |  |  | 49.9 (5.2) |  |  |  |
| 3. SMC | 2,631 (15.5%) | 19,347 | 4,462 (9.2%) | 48.9 (8.9) | All | GE (2,596) | None |
|  | (409/2,222) |  | (410/4,052) | 47.2 (7.0)/ | Korean | Hologic (495) |  |
|  |  |  |  | 49.2 (9.1) |  | Lorad (1,371) |  |
| 4. BMC | 752 (5.5%) | 5,674 | 754 (5.6%) | 53.6 (13.2) | All | Hologic (754) | None |
|  | (41/711) |  | (42/712) | 72.1 (6.0)/ | Korean |  |  |
|  |  |  |  | 52.5 (12.6) |  |  |  |
| 5. IPH | 148 (23.0%) | 402 | 148 (23.0%) | 52.0 (12.7) | All | Hologic (148) | None |
|  | (34/114) |  | (34/114) | 56.8 (14.0)/ | Korean |  |  |
|  |  |  |  | 50.6 (12.1) |  |  |  |
| Total | 12,312 (20.2%) | 67,749 | 13973 (17.5%) | 52.8 (9.8) | All | GE (6,982) | 14,031 |
|  | (2,445/9,687) |  | (2,447/11,526) | 51.3 (10.0) / | Korean | Hologic (5,620) |  |
|  |  |  |  | 53.2 (9.7) |  | Lorad (1,371) |  |
| Replication Study |  |  |  |  |  |  |  |
| EMBED<br>(20%) | 4,696 (1.9%) | 222,880 | 5,022 (1.91%) | 58.8 (11.9) | All | Hologic (4,037) | None |
|  | (89/4,607) |  | (96/4,926) | 61.1 (15.1) / | White* |  |  |
|  |  |  |  | 58.7 (11.8) |  | GE (985) |  |

Abbreviations : NCC – National Cancer Center, Korea; AMC- Seoul Asan Medical Center, Korea; SMC – Samsung Seoul Medical Center, Korea; BMC – Seoul Boramae Medical Center, Korea; IPH – Inje University Ilsan Paik Hospital, Korea; EMBED – Emory Breast Imaging Dataset, United States
\*White women’s data were used

Selected images underwent preprocessing, including normalization to reduce inter-vendor variability in image feature variability. Changes in image feature differences before and after the normalization visualized by UMAP is shown in fig. 4.

**Fig 4.**
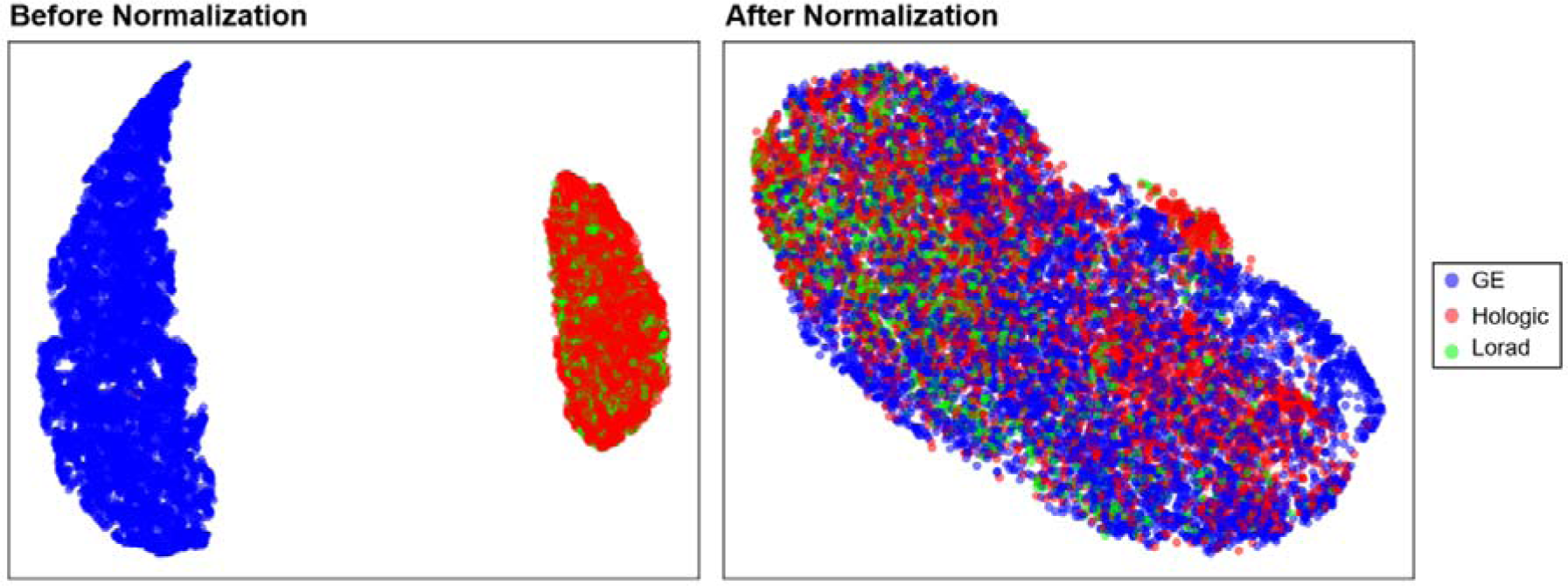
Image feature distribution before (left) and after (right) the normalization between vendors. Uniform Manifold Approximation and Projection (UMAP), a multifactor dimensionality reduction method was used for visualizing the high-dimension features between vendors.

### 3.2 MBD comparison before and after skin removal

Importantly, we discovered a significant impact on pMBD (percent of MBD to total breast area) metrics when skin was removed from the breast area during semantic segmentation (fig. 5). After skin removal, the pMBD metrics shifted notably: for cumulus, altocumulus, and cirrocumulus, the mean values changed from 20.3%, 6.7%, and 0.84% to 16.5%, 5.1%, and 0.34%, respectively. This highlights the potential for cirrocumulus to be particularly overestimated without proper skin exclusion.

**Fig. 5.**
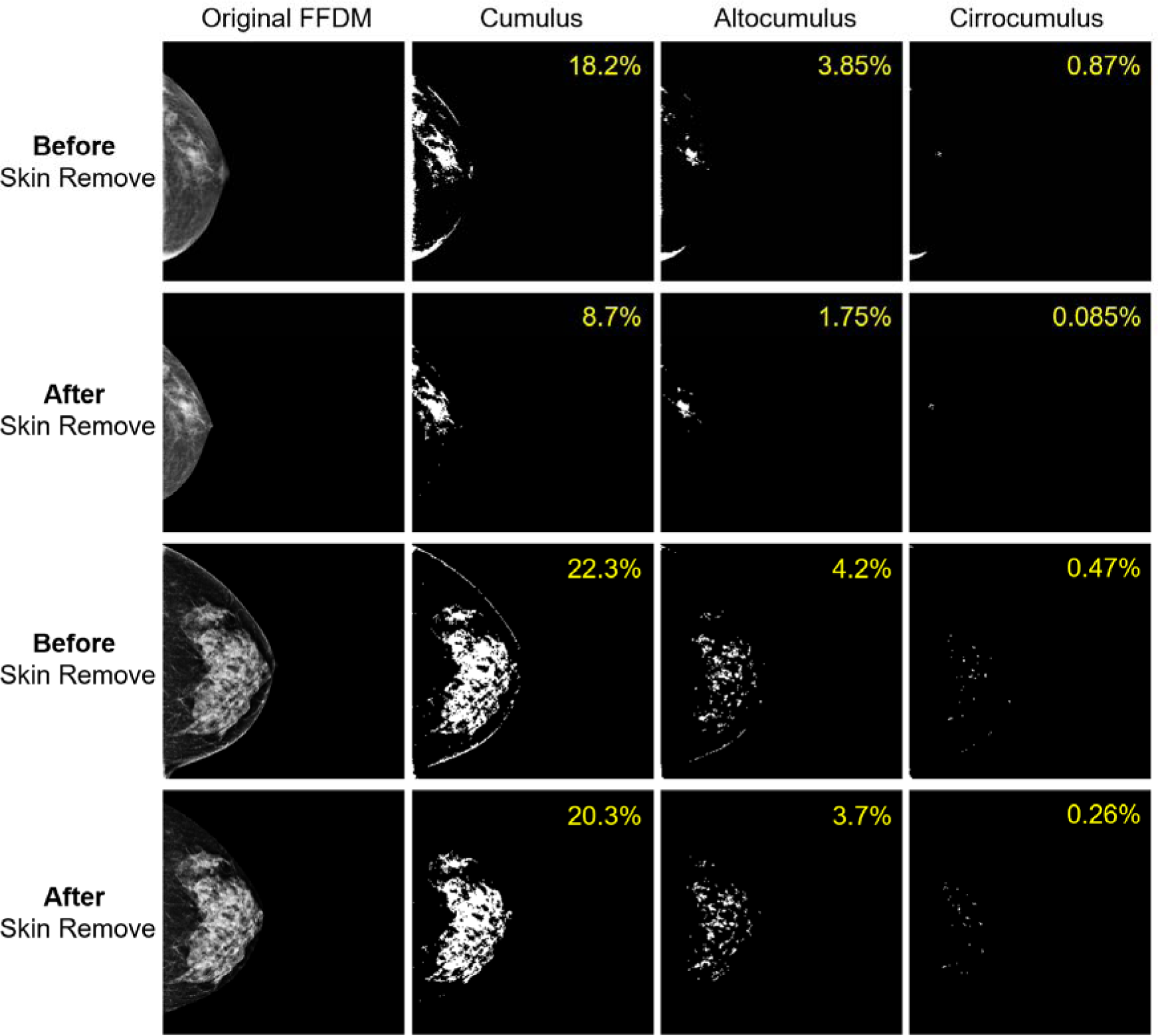
Multi-level MBD measurements before and after the removal of skin from breast area. Figure shows examples of two FFDM images. First and third row shows MBD of FFDM images before skin removal, and second and fourth row shows MBD of skin removed version of corresponding images. pMBD values are shown in upper lower area of each MBD masks.

### 3.3 Automated multi-level MBD measurement and FS from DL

The automated multi-level MBD measurements correlated strongly with expert evaluations. Spearman’s correlation coefficients were 0.956, 0.945, and 0.87 for cumulus, altocumulus, and cirrocumulus (Supplementary Table 1). The adjusted pMBD measurements demonstrated significant differences between case and control groups in the independent test dataset. When benign tumors were included in the analysis (measurement of the contralateral side of the tumor), the pMBD distribution of the benign tumors were located in midway between case and control images (fig. 6A- 6C). The differences in pMBD distributions were more pronounced for altocumulus and cirrocumulus than for cumulus, the conventional MBD measurement.

**Fig. 6.**
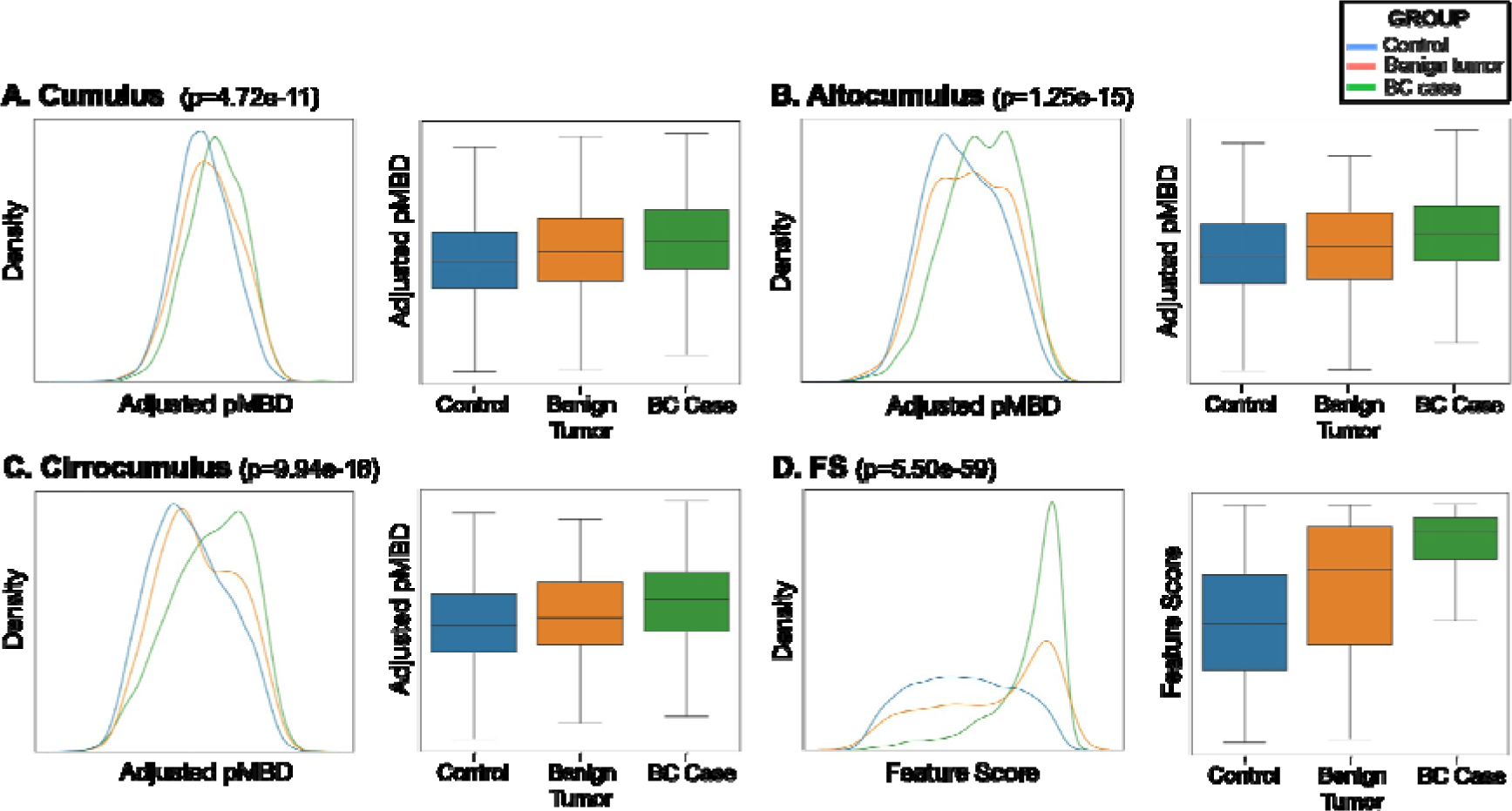
Distribution of adjusted multi-level percent MBD (pMBD) (A-C) and feature scores (FS) derived from deep learning (DL) (D). Two-tailed two-sample t-tests were applied for comparing control, benign tumor and BC case images. *P*-values from comparison between control and BC cases are shown. (*P*-values of comparison between control and benign tumors, benign tumors and BC cases are shown in Supplementary Table 2.)

When it comes to distinguishing between BC case and control groups, the FS derived from DL exhibited a marked difference. The FS for benign tumors displayed a broader distribution, positioning it between the BC cases and controls (fig. 6D).

### 3.4 Predictive values of individual, and combined cirrocumulus and FS

When adjusted for age and total breast area, the pMBD measured alone offered a moderate Area Under the Receiver Operating Characteristic Curve (AUROC) ranging from 0.63 for cumulus to 0.67 for all combined measures (fig. 7). This suggests a moderate predictive power for each individual MBD measure, with a slight improvement when focusing on alto- and cirrocumulus measures. In a multivariate logistic regression model, the “odds per unadjusted standard deviation (OPERA)” (Hopper, 2015) demonstrated that OPERA of cumulus is attenuated from 1.71 (univariate) to 1.21 (adjusted by cirrocumulus), whereas the cirrocumulus is slightly attenuated from 1.75 to 1.53 (Supplementary Table 3). The FS from DL outperformed pMBD with an AUROC value of 0.82, alongside an Area Under the Precision-Recall Curve (AUPRC) of 0.51, sensitivity of 0.91, and precision of 0.26 using the test set of Korean data (fig. 7). Adding MBD information to FS did not improve the AUROC.

**Fig. 7.**
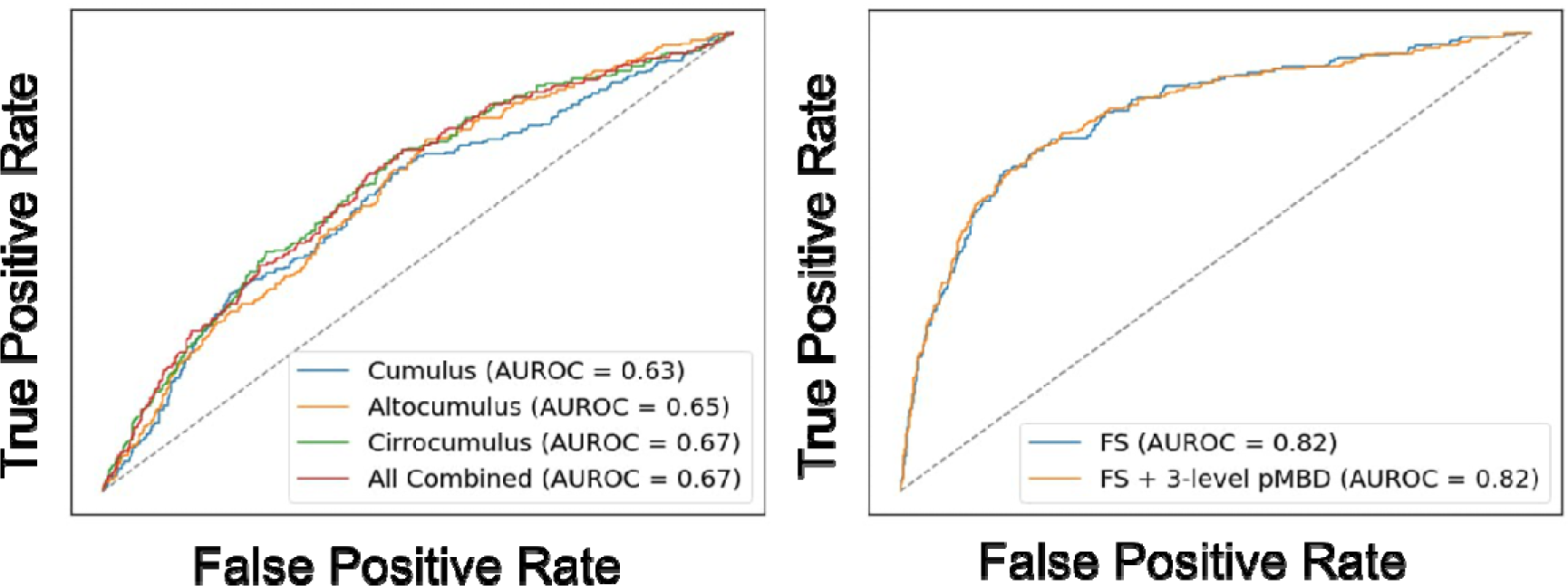
Predictive values of each and combined pMBD and FS from DL for BC. Area under the ROC curve (AUROC) for three pMBD measures (left), FS and FS combined with three pMBD measures (right).

FS and adjusted pMBD showed moderate positive correlation (r=0.35, *p*<0.001). Interestingly, an increase in adjusted pMBD correlated linearly with an increase in the BC prevalence in higher FS groups, but not in the mid- or lower FS groups (fig. 8).

**Fig. 8.**
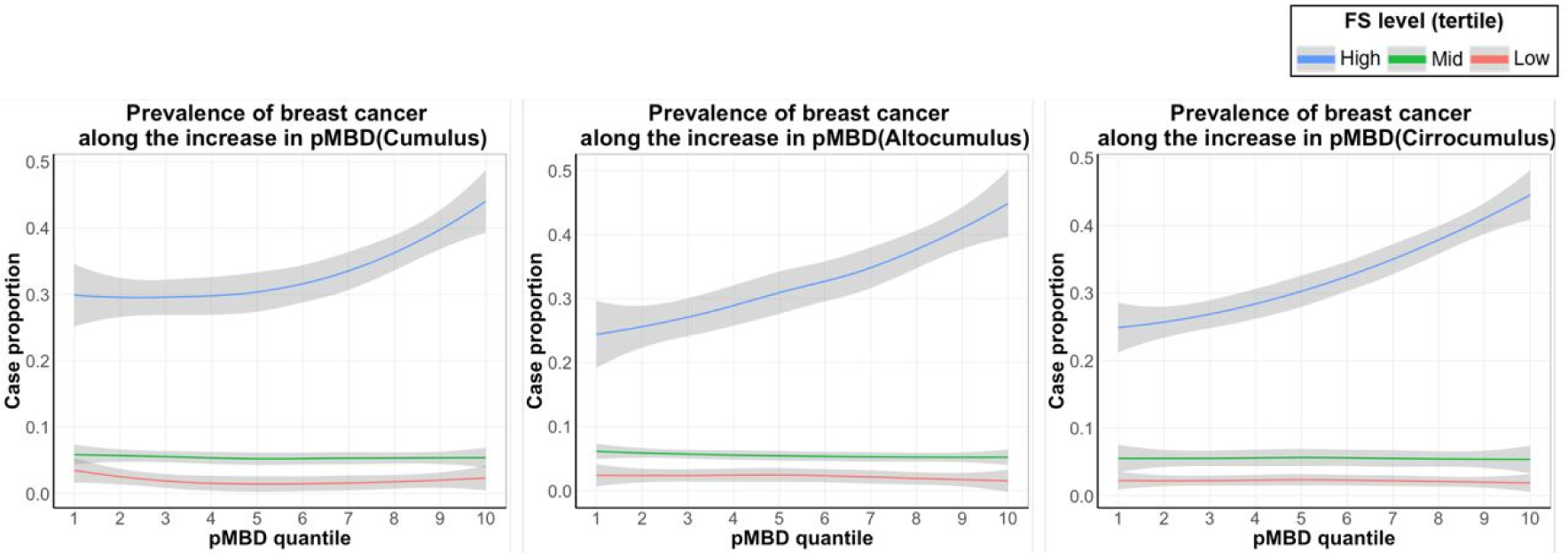
Trend of the prevalence of BC along the increase in the three levels of pMBD, stratified by FS tertile group. Left: Cumulus, Middle: Altocumulus, Right: Cirrocumulus. Each line represents the FS tertile group (Blue: Highest FS tertile group, Green: Intermediate FS tertile group, Red: Lowest FS tertile group)

While FS excels in predicting BC risk, when both FS and pMBD measures were combined, the results were noteworthy (fig. 9). The subgroup that had the higher tertile for both FS and cirrocumulus (“double higher”) had an odds ratio (OR) of 10.20 (95% CIs 7.37-14.11) relative to the mid tertile FS group in Korean women (fig. 9A). The estimation was made by using 20% of independent test dataset only. For all cases of the US White women (EMBED), the same measure showed OR=5.67 (95% CIs 3.10-10.37) (fig. 9B), but further discrimination by MBD was not evident or even reversed for generic MBD level. When we focused on the screen-detected cases (US only), the OR increased to 7.09 (95% CIs 2.82-17.83) with distinct risk gradation by MBD level in upper-tertile FS (fig. 9C). Notably, in subset of screen-detected cases, only cirrocumulus showed additional discriminative power to FS in upper-tertile FS. This synergistic effects between FS and cirrocumulus got stronger when we selected higher FS (upper-10%), OR gets as high as 19.05 (13.31-27.26) for Korean (FS upper-10% and cirrocumulus upper-tertile), and 15.91 (6.12-41.38) for the same subgroup applied to screen-detected cases in US (Supplementary fig. 1A and 1B). Same analysis applying more strict pMBD of top-10% further increased risk, OR=23.59 (14.65-37-99), which was only conducted in the test dataset of Korean due to the decreases in sample size (Supplementary fig. 1C). Notably, the MBD information, particularly cirrocumulus, consistently provided additional discriminative power to FS no matter what the threshold might be, but this gain was only observed for women with higher FS.

**Fig. 9.**
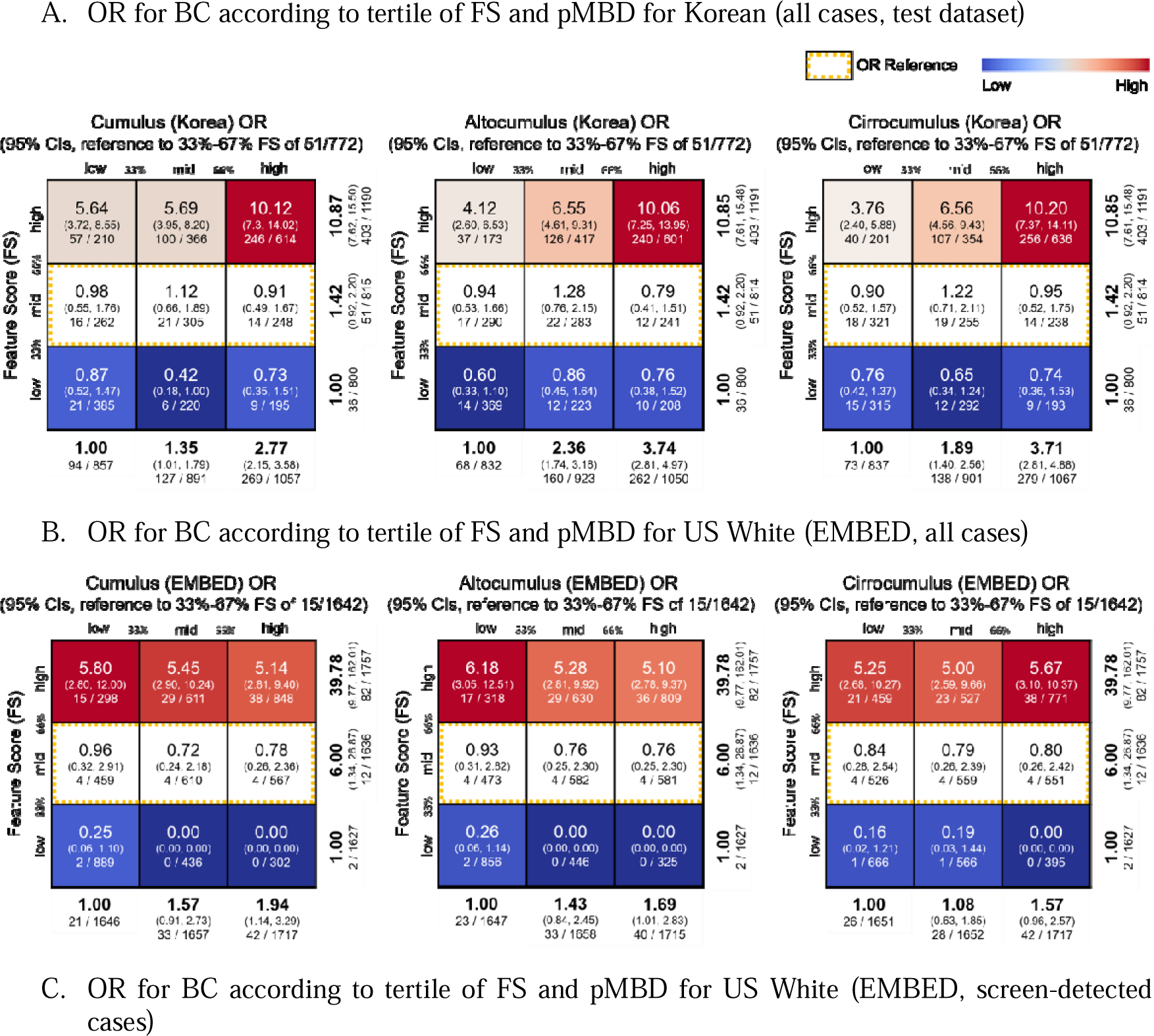
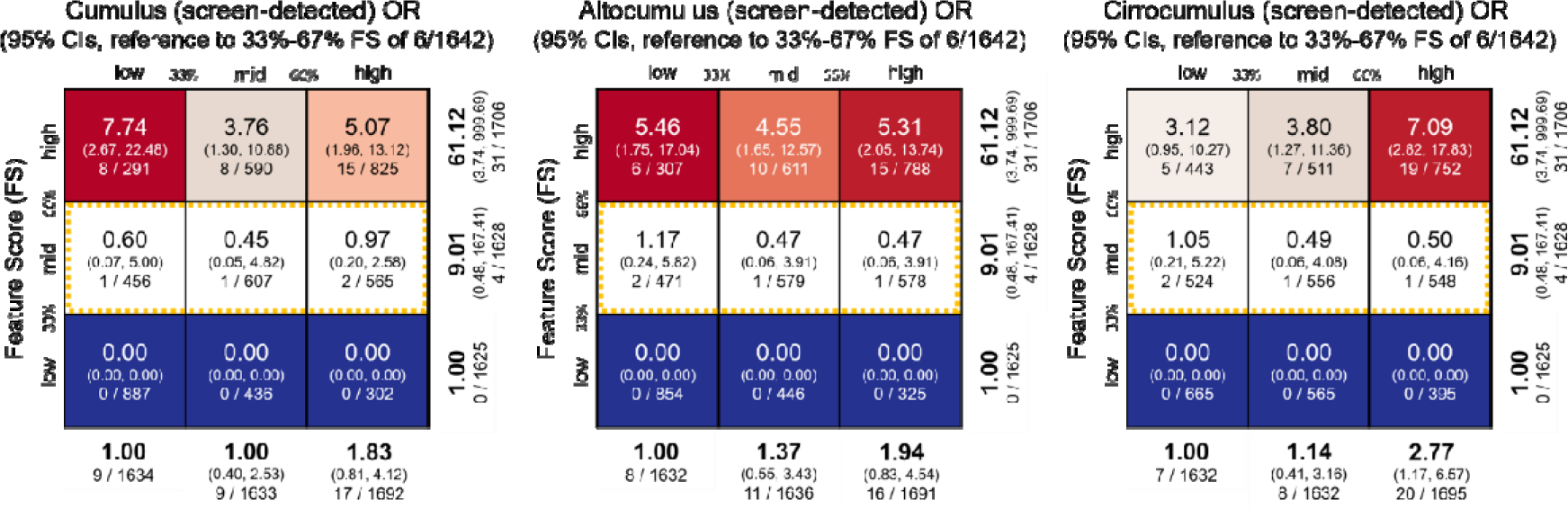
Odds ratio (OR) of BC according to tertile of FS and adjusted pMBD, reference to mid tertile FS group. (A) All cases of Korean test dataset (20% of all data, upper), (B) All cases of US (EMBED) data (middle), and (C) Screen-detected cases from US EMBED. Within each cell, OR is shown in top, followed by 95% CIs. Numbers of cases and controls are described in the bottom of cells. Marginal ORs for each row and columns are also described.

### 3.5 Deciphering the risk features from DL

When we visualized the FS importance using guided Grad-CAM, and juxtaposed the feature importance with the original, and multi-level MBD images, the features generally captured higher-intensity MBD measures as exhibited in fig. 10. When we only focused on the “highly important” patterns with feature importance weighting >0.9, they captured patterns predominantly from cirrocumulus and some part of altocumulus (example #1 and #2 in fig. 10). However, as shown in the #3 individual in fig. 10, some examples also indicate the FS are also using complex textures beyond cirrocumulus as well.

**Fig. 10.**
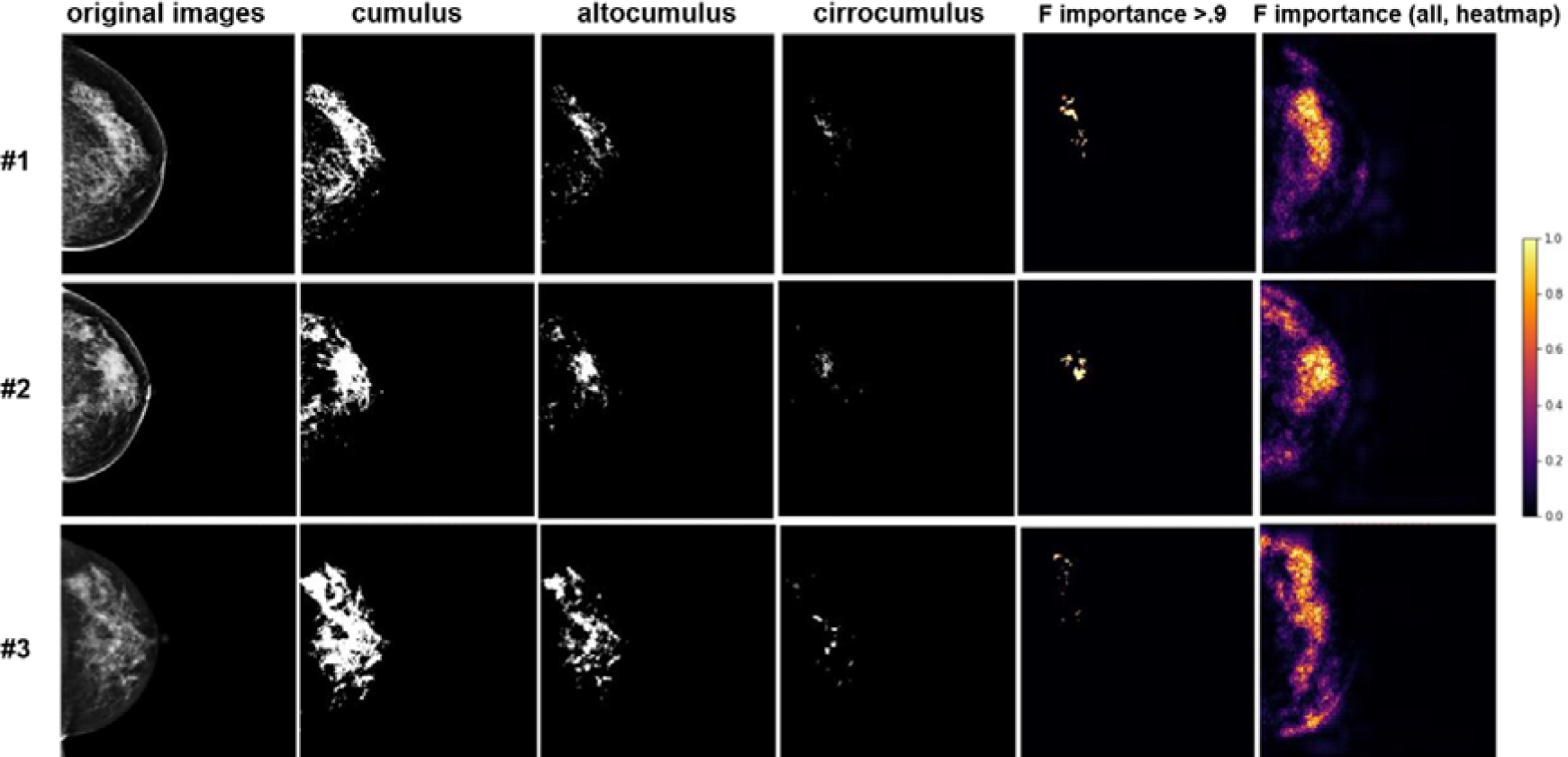
Examples of the feature importance of “double-higher” derived from DL in comparison with automatically measured multi-level MBD. Feature importance (F importance) was estimated using Guided Grad-CAM. The last column shows heatmap of all features, and the penultimate column only shows the most important features (F importance > 0.9). Three images are randomly selected from individuals in the “double-higher” group in fig. 9.

## 4. Discussion

Our study suggests that FFDM can be used not just for diagnosis but also for automated risk prediction. In this study, we’ve demonstrated that FS derived from DL show significant promise for risk prediction in breast cancer, outperforming traditional models or metrics such as MBD. This aligns with and strengthens the existing evidence supporting the utility of DL in predicting the risk of BC (Yala et al., 2022). Importantly, our analysis goes a step further to suggest that FS aren’t just statistically significant indicators, but may actually represent complex textural and morphological features in mammographic images. These are features that may not be captured fully by existing metrics like MBD, thus adding a new layer of information for risk prediction.

Yet, while the FS show excellent predictive potential, they may not be fully self-sufficient. The predictive value of FS increases according to MBD, especially in top tier of FS tertile (fig. 8). We believe this particular relationship between FS and MBD may explain why linear models adding MBD to FS does not enhance the predictive performance substantially (fig. 7). Our findings show that an integrated model that stratifies both FS and MBD better detect a subgroup individuals with substantially increased risk for BC. We’ve also shown that by combining FS with MBD, particularly cirrocumulus, the brightest density, high-risk subgroups for BC can be identified.

### 4.1 Quality control and image preprocessing in our study

In our study, we implemented rigorous quality control measures, including vendor-specific normalization. UMAP analysis confirmed the effective reduction of inter-vendor variability by our approach (fig. 4). Importantly, we introduced a skin-removal technique, an often-overlooked step in conventional automated segmentation. Our results indicate that the inclusion of skin erroneously inflates higher-intensity MBD measures, particularly cirrocumulus, as shown in fig. 5. The skin area is regarded as dense area for all levels of MBD measurement, with the relative influences particularly larger for cirrocumulus. As depicted in fig. 5, the presence of skin can inflate the cirrocumulus measurement by more than two-folds, in average from 0.34% to 0.87%. By contrast, skin removal was unnecessary for DL-based feature score estimation, as skin presence didn’t significantly impact the risk prediction.

### 4.2 Automation of high-intensity MBD measurement

Our work corroborates that more granular MBD measures such as altocumulus and cirrocumulus can be more informative than the traditional cumulus measures. The predictive value as assessed by the AUROC was higher for these granular MBD categories. High-intensity signals like cirrocumulus have limited their broader adoption due to measurement challenges. Our automated system offers a robust solution, correlating closely with expert evaluations with Spearman’s coefficients of 0.87 for cirrocumulus. Given that the correlations for expert’s measurement is 0.98, 0.93, and 0.80 for cumulus, alto- and cirrocumulus (Nguyen et al., 2015), our automated measurements stand as a reliable alternative. The MIDAS was developed to normalize two major vendors with strict image pre-processing including automatic skin and artifacts removal. We already showed that our new pre-processing methods improve the automatic measurements particularly for cirrocumulus. (fig. 5). Although cirrocumulus is a suspected key risk factor within MBD (Nguyen et al., 2020; Nguyen et al., 2021), more data is needed to become a standard practice. Our method could expedite its mainstream acceptance.

### 4.3 FS from DL combined with cirrocumulus: to identify high-risk individuals

DL-derived FS outperformed MBD metrics in predictive value. In linear models, such as logistic regression, MBD’s additional contribution to FS was negligible. Interestingly, in all MBD tertiles, increase in FS was strongly associated with the prevalence of BC (fig. 9). Yet, MBD’s significant predictive contribution is only evident in the highest FS tertile. The cirrocumulus show more discriminative to FS, than cumulus or altocumulus. Leveraging both the cirrocumulus level and high FS, our approach identifies a subgroup with significantly increased odds ratios (OR) for breast cancer – 10.2 for Koreans and 5.67 for US Whites. Given the definition of “high-risk” individuals by ACS, >20-24% life-time BC risk (Smith et al., 2019), we believe the “double-higher” group individuals can become novel candidates for specialized high-risk screening, although our study does not directly provide life-time risks. Despite the constraints from the case-control study designs, we believe our findings have the potential to transform BC prevention. First, our findings are in alignment with the definition of high-risk, life-time risk >20-24% for BC, if we apply the baseline population life-time BC risk of 5% in Korean women (National Central Cancer Registry, 2023), or 10% of US White ((IARC), 2021). Note that we derived the thresholds only from the control data of two populations and the cut-offs are not affected by the data of cases. Second, our method idenfies intended proportion of subgroup at risk, e.g. ∼10% of population by upper-tertile FS, or 3% by upper-10%, suitable for 2-step approaches in mass screening programs. Third, our approaches soly relies on mammography and fully automated measurement, so that ours provide unprecedent compliance in detecting high-risk individuals. The information of the BC risk from mammography can be used with other important risk factors or stand-alone, according to the study setting.

### 4.4 Consistency and reliability of the high-risk subgroup

Our findings generally show agreement between two populations but the OR is higher in Koreans. The differences in OR might arise from the fact that we first trained the FS algorithms using Korean women’s data, then transferred them to US women. However, we estimated FS and MBD for Korean and the US women using independent dataset, and it is not likely that the differences arise from the information leakage. Additionally, the Korean and US data differ in their study designs. While the EMBED dataset is a mixture of cohort and case-control designs, the Korean dataset encompasses higher proportion of cases from case-control designs. The OR could have exaggerated true risk, as the higher proportion of BC cases in Korean data represent the characteristics of high-risk individuals. An intriguing observation was the pronounced relevance of cirrocumulus over other MBD measures, especially when dividing the EMBED dataset into screen-detected and other types (fig. 9C and Supplementary fig. 1B). The subset with screen-detected cancers in EMBED exhibited a clear risk gradient by cirrocumulus, but less or even reversed patterns for altocumulus or cumulus level. While results from screen-detected cases have more significant implications, the broader confidence intervals by decreased sample size may require further validation.

### 4.5 Demystifying the feature scores from deep learning

We applied guided Grad-CAM, an “explainable AI (XAI)” methods, to understand the feature importance (fig. 10). Our findings suggest that the FS generally agrees with the alto- and cirrocumulus area using visual inspection. Given that the DL makes high-dimensional inferrences, it is not directly understandable what shapes or complex textures of the FFDM are used to predict the risk of BC. Our study along with the previous studies using DL (Yala et al., 2022) demonstrates that the mammography does include invaluable predictive information as well as for diagnostics. Our study adds new insights that the FS mainly utilize the information of dense area, particularly alto- or cirrocumulus.

### 4.6 “Case Images” and “Control Images” in this Study

We followed the convention of defining cases and controls widely used in MBD studies (Boyd et al., 2007); using the contralateral side of the cancer lesion as “case image”, and a randomly selected side as “control”. The tumor lesions are dense with frequent calcification. In estimating future risk, the consequent changes in MBD by tumors have been excluded to improve predictive accuracy. We believe the same rationale is applicable to DL studies and multi-level MBD analysis. We admit that the longitudinal definition of cases, i.e., the ipsilateral side of mammography that will develop cancerous lesion in future, might be better. In our study, the case-control settings made the use of conventional definition of “case images” inevitable. Our findings may need to be corroborated in longitudinal data and subsequent case definition.

### 4.7 Limitations of the Study

We admit that our study has several limitations. First, our study established its methods primarily using case-control study design data. The case-control nature of our dataset precluded us from calculating accurate risk metrics like the C-index or 10-year incidence risk. Given the definition of the high-risk by ACS, the increase in life-time risk more than 20-24% confer particular meaning in preventive strategies. Our findings were confirmed in longitudinal data of screen-detected cases, but a large-scale study longitudinal data may be needed to fully corroborate our findings. Second, our findings that the sugroups of “double-higher” have substantially increased risk of BC need to be validated in broader populations. Third, we included FFDM images from two major vendors, GE and Hologics, but not for other vendors. To utilize the same method for the images generated by other vendors additional preprocessing methods should be established. However, we believe our normalization methods can be used for the multi-vendor data consiting of the major vendors only. Four, our study only included 2-dimensional FFDM. Considering the recent increase in 3D tomographic mammography, 3D module also need to be developed, as the accessibility to those database is acquired. Lastly, we did not consider other important risk factors of BC, particularly the genetic risk factors and family history of BC. Many studies, including ours, large cohort studies do not collect mammographic images, and FFDM databases does not have genetic and epidemiologic information. While we anticipate that incorporating other well-established risk factors could enhance the model’s overall predictive accuracy, the practical utility of our approach lies in its reliance solely on mammography data. This is particularly relevant when considering the limited accessibility of detailed genetic, clinical, or epidemiological information in many settings.

## 5. Conclusion

In conclusion, despite the limitations outlined earlier, our study affirms the unique predictive value offered by FFDM. We introduce an automated method for quantifying multi-level MBD, a tool that can be invaluable in related research. Additionally, we demonstrate the efficacy of identifying the “double-higher”, a synergistic approach that leverages both DL and the concept of “cirrocumulus” in identifying a high-risk subgroup of women solely through mammography. The broader applicability and usefulness of our findings will ultimately depend on their effect size and efficiency, which will be further assessed in larger-scale validation studies.

### Accessibility of data and source codes

The Korean data for mammographic images are not available for open access. The EMBED data requires a process for online data access. The source code for MIDAS analysis are freely accessible and can be found in https://github.com/ghbd-snu/MIDAS-MD.

## Data Availability

The Korean data for mammographic images are not available for open access. The EMBED data requires a process for online data access

## Acknowledgement

We would like to express our gratitude to Dr. Hari Trivedi for generously sharing data from EMBED, also to colleagues in SMC and NCC for sharing data.

## Abbreviations

BC: breast cancer
MBD: mammographic breast density
DL: deep learning
FS: feature score
FFDM: full-field digital mammography
DICOM: digital imaging and communications in medicine
XAI: explainable artificial intelligence
DCIS: ductal carcinoma in situ

