## Supplementary material for "Artificial-Intelligence Powered Identification of High-Risk Breast Cancer Subgroups Using Mammography: A Multicenter Study Integrating Automated Brightest Density Measures with Deep Learning Metrics": Supplmentary information

Supplementary Methods

### **Supplementary Methods 1**

#### **Focal Tversky Loss**

For semantic segmentation, the Dice score coefficient (DSC) is widely used to assess segmentation performance. The 2-class DSC variant for class $c$ is expressed in equation (1), where $g_{ic}\in\left\{ 0,1 \right\}$ and $p_{ic}\in\left[ 0,1 \right]$ represent the ground truth label and predicted label, respectively. The total number of pixels in an image is denoted by $N$. The $\epsilon$ provides numerical stability to prevent division by zero. The linear Dice loss (DL) is defined a minimization of the overlap between the prediction and ground truth (Milletari et al., 2016).

$$DSC_{c}=\frac{\sum_{i=1}^{N} p_{ic}g_{ic}+\epsilon}{\sum_{i=1}^{N} p_{ic}+g_{ic}+\epsilon} (1)$$

One of the limitations of the DSC is that it equally weights false positive (FP) and false negative (FN) detections. For efficient segmentation of multi-level MBD, we used Tversky index (TI) to quantify segmentation quality. The TI is a generalization of the DSC which allows for flexibility in balancing FP and FNs (equation (2)) :

$$TI_{c}=\frac{\sum_{i=1}^{N} p_{ic}g_{ic}+\epsilon}{\sum_{i=1}^{N} p_{ic}g_{ic}+\alpha\sum_{i=1}^{N} p_{i\bar{c}}g_{ic}+\beta\sum_{i=1}^{N} {p_{ic}g}_{i\bar{c}}+\epsilon} (2)$$

where, $p_{ic}$ is the probability that pixel $i$ is of the lesion class $c$ and $p_{i\bar{c}}$ is the probability pixel $i$ is of the non-lesion class $\bar{c}$. The same is true for $g_{ic}$ and $g_{i\bar{c}}$, respectively. Hyperparameters $\alpha$ and $\beta$ can be tuned to shift the emphasis to false positive and false negative detections. The Tversky index is adapted to a loss function (TL) by minimizing $\sum_{c} 1-TI_{c}$ (Hashemi et al., 2018). In case of $\alpha=\beta=0.5$, TI simplifies to the DSC. If $\alpha>0.5,$TI weights more emphasis on minimizing FN predictions, while setting $\beta$ larger than 0.5 weights more emphasis on minimizing FP predictions.

In practice, DL struggles to segment small region of interest (RoI) as they do not contribute to the loss significantly. To mitigate this problem, the authors of improved attention U-Net paper (Abraham and Khan, 2019) proposed the focal Tversky loss function (FTL), which is parameterized by hyperparameter $\gamma$ to control between easy background and hard RoI training examples. The focal parameter exponentiates the cross-entropy loss to focus on hard classes detected with lower probability (Lin et al., 2017). FTL is defined as (equation (3)) :

$$FTL_{c}=\sum_{c} \left( 1-TI_{c} \right)^{1/\gamma} (3)$$

When $\gamma>1$, the loss function focuses more on less accurate predictions that have been misclassified. Authors observed the best performance with $\gamma=\frac{4}{3}$. However, they found that using FTL as loss function for all layers over-suppressed FTL when the model is close to converge. So they recommended to train intermediate layers with FTL but supervised the last layer with TL to provide a strong signal and mitigate sub-optimal convergence. In this paper, we adapted recommended architecture of improved attention U-Net, but changed values of $\alpha$ and $\beta$ to accurately segment dense regions corresponding to cumulus, altocumulus, and cirrocumulus.

#### **Network Architecture**

The improved attention U-Net (Abraham and Khan, 2019) is based on U-Net (Ronneberger et al., 2015), which is composed of a contracting path to extract locality features and an expansive path, to resample the image maps to combine high-resolution local features with low-resolution global features and encourage more semantically meaningful outputs.

However, at the deepest stage of encoding where the network has the richest possible feature representation, spatial details tend to get lost in the high-level output maps. This makes it difficult to reduce false detections for small objects that show large shape variability (Oktay et al., 2018). So the attention gates (AG) is added to vanilla U-Net architecture to identify relevant spatial information from low-level feature maps and propagate it to the decoding stage. With additive attention gate, input features are scaled with attention coefficients to propagate relevant features to the decoding layer output. The coarser gating signal provides a contextual information while spatial regions from the input features provide locality information. Feature map is then resampled by bilinear interpolation. Details for attention-gated U-Net are explained in its original paper (Oktay et al., 2018).

#### **Segmentation Model Development**

The fully-automated multi-level MBD segmentation model was based on improved attention U-Net described above. As the model receives both input image and corresponding ground truth mask, images with experts’ manual measure of multi-level MBD (cumulus, altocumulus, cirrocumulus) were used for semantic segmentation model development. We acquired three ground truth (MBD) masks (cumulus, altocumulus, cirrocumulus) for each image, by retaining pixels with higher intensity than these experts’ manual measure of multi-level MBD threshold values. Using these image – MBD mask pairs, we trained segmentation model to predict multi-level MBD masks. Keras API with tensorflow was used for model deployment. Dataset was randomly splitted into 8 : 1 : 1 as training : validation : test set, stratifying the vendor. Since three levels of MBD have substantially different areas for same input image, we trained each segmentation model with different hyperparameter $\alpha$ for TL to find optimal balance between FP and FN. We used $\alpha$ value of 0.1, 0.2, …, 0.9, and choose $\alpha$ which showed maximum DSC between ground truth in test images. Hyperparameters and test DSC of multi-level MBD segmentation networks are shown in Supplementary Table 1. All the models were trained with Adam optimizer with initial learning rate 0.0003 and momentum value of 0.9.

### **Supplementary Methods 2**

#### **Binary Classification Model Development**

Dataset was splitted into 7:1:2 as training : validation : test set, stratifying the clinical centers, vendors with preserving overall case-control ratio. We trained DenseNet 121(Huang et al., 2017). Model was trained with Ranger optimizer(Wright, 2019) (initial learning rate 1e-3) since this optimizer confers advantages in convergence speed, generalization, and stability over other optimizers. For augmentation, we applied random flips and contrast changes, to avoid the distortion or omission of risk features. To mitigate imbanace between case and control, we applied the case-control ratio as the weight in calculating loss function of binary cross entropy. We conducted model training by 100 epochs and saved the model with highest recall in validating dataset, to prioritize the detection of high-risk individuals. The risk feature score (FS) was calculated by applying sigmoid function to the output of trained CNN for binary classification.

#### **Fine tuning for EMBED**

The binary classification model was initially developed solely by korean dataset (13,973 images). To estimate FS for EMBED, we applied fine-tuning methodology to alleviate potential variations attributed to ethnicity. In this process, none of the model layers were held fixed, and we fine tuned the model with EMBED dataset. The training parameters and specifics remained consistent with the previously described methodology.

### **Supplementary Methods 3**

#### **pMBD adjustment**

pMBD was calculated as percentage of each multi-level MBD by total breast area. As there were multiple studies describing relationship between age, body mass index (BMI) and pMBD, we adjusted pMBD measurements. We fitted linear regression model, which featured age and total breast area as independent variables, while log-transformed pMBD served as the dependent variable. Given the availability of BMI measurements for a subset of individuals, we leveraged the total breast area as a surrogate for BMI. This decision was informed by the verified linear correspondence existing between BMI and total breast area. (R^2^ 0.3376, *p* < 0.001). The residuals were subsequently calculated from fitted linear regression model, and we applied Yeo-Johnson transformation to residuals (Weisberg, 2001). The resultant transformed values (adjusted multi-level pMBD measurements) were used for breast cancer risk prediction.

We conducted additional version of adjusting MBD by leveraging the non-dense area (total area absracted by cumulus area) as a surrogate for BMI. These results are shown in Supplementary Figure 3.

Supplementary Results

#### **Supplementary Table 1.** Correlation between MIDAS and experts’ measurement for (as Spearman’s correlation coefficients). Dice score between experts’ measurement and fully-automated measurement are shown in second column. Last column shows selected optimal hyperparameter $\alpha$ for semantic segmentation network.

|  | **Correlation Coefficient** | **Dice Score** | **Hyperparameter** $\boldsymbol{\alpha}$ **for Tversky Loss** |
| --- | --- | --- | --- |
| **Cumulus** | 0.956 | 0.915 | 0.6 |
| **Altocumulus** | 0.945 | 0.815 | 0.5 |
| **Cirrocumulus** | 0.87 | 0.682 | 0.5 |

#### **Supplementary Table 2**. Two-tailed two-sample t-test *p*-values in comparison of adjusted multi-level pMBD and FS between control, benign tumor and BC cases.

|  | **BC case vs. Control** | **BC case vs. Benign tumor** | **Control vs. Benign tumor** |
| --- | --- | --- | --- |
| **Cumulus** | 4.72e-11 | 9.8 e-07 | 1.18e-03 |
| **Altocumulus** | 1.25e-15 | 1.63e-04 | 8.84e-07 |
| **Cirrocumulus** | 9.94e-18 | 4.64e-06 | 1.13e-07 |
| **FS** | 5.50e-59 | 4.54e-22 | 1.97e-26 |

#### **Supplementary Table 3.** OPERA scores for normalized age, total area, cumulus, and cirrocumulus

| **Factor** | **OPERA** | | $\boldsymbol{p}$ | | **AIC** |
| --- | --- | --- | --- | --- | --- |
| Cumulus | AGE (Normalized) | 1.07 | 0.00146 | * | 12355 |
|  | Total area (Normalized) | 1.49 | < 2e-16 | *** |  |
|  | Cumulus (Normalized) | 1.71 | < 2e-16 | *** |  |
| Cirrocumulus | AGE (Normalized) | 1.00 | 0.793 |  | 12255 |
|  | Total area (Normalized) | 1.31 | <2e-16 | *** |  |
|  | Cirrocumulus (Normalized) | 1.75 | <2e-16 | *** |  |
| Cumulus + Cirrocumulus | AGE (Normalized) | 1.05 | 0.11 | . | 12235 |
|  | Total area (Normalized) | 1.36 | < 2e-16 | *** |  |
|  | Cumulus (Normalized) | 1.21 | 2.45e-06 | *** |  |
|  | Cirrocumulus (Normalized) | 1.53 | < 2e-16 | *** |  |

* Abbreviation : OPERA, odds per adjusted standard deviation

Supplementary Figures

### **Supplementary Fig. 1.**

#### **A.** OR for BC according to upper-10% of FS (lower 10%, mid 80%, upper 10%) and adjusted pMBD tertile for Korean (all cases, test dataset) , reference to mid tertile FS.

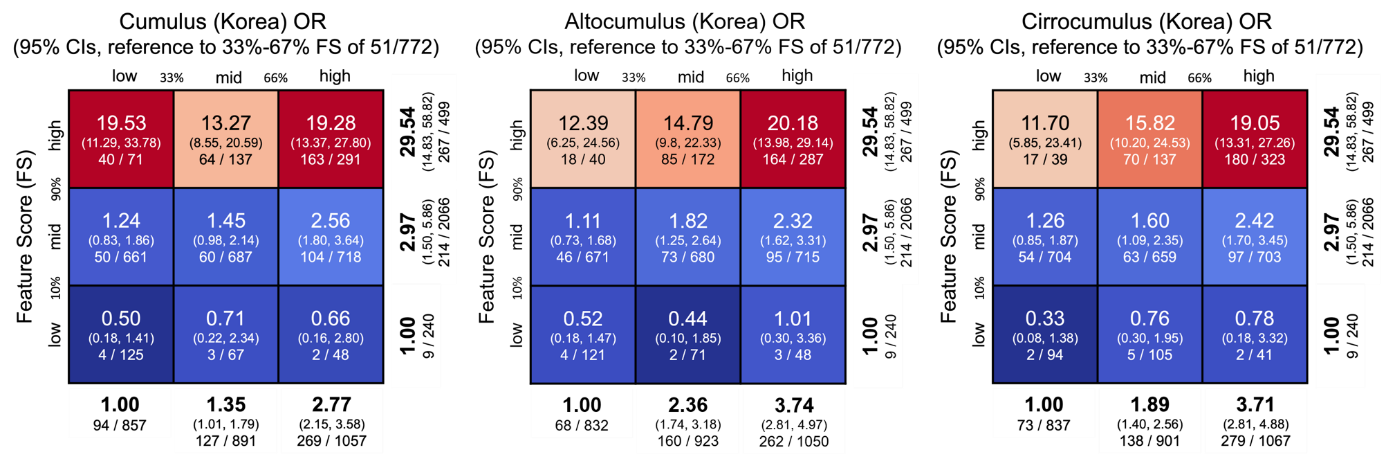

#### **B.** OR for BC according to upper-10% of FS (lower 10%, mid 80%, upper 10%) and adjusted pMBD tertile for US White (screen-detected cases) , reference to mid tertile FS.

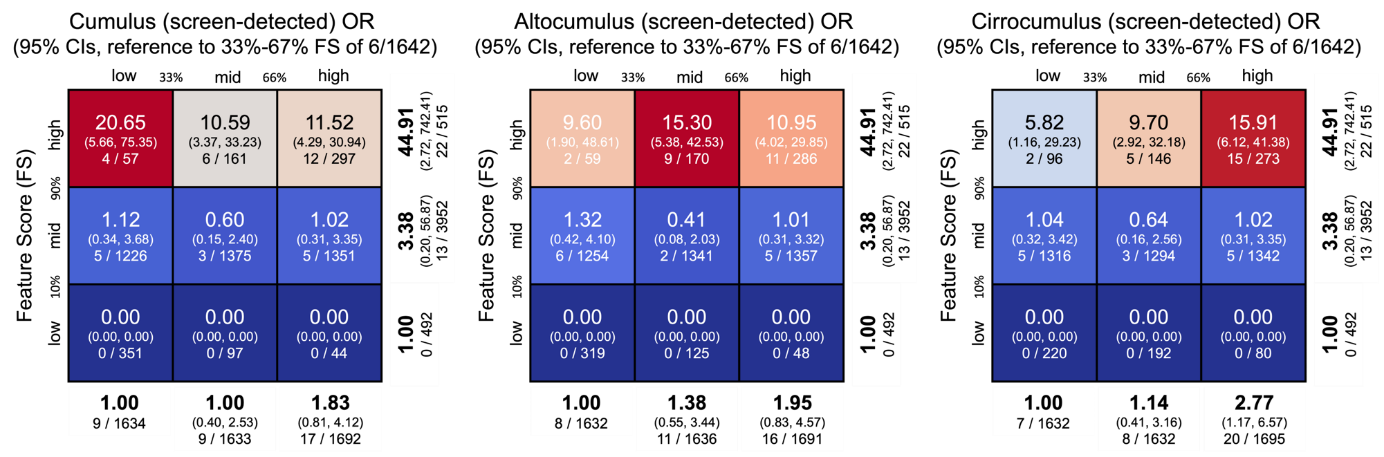

#### **C**. OR for BC according to upper-10% of FS (lower 10%, mid 80%, upper 10%) and upper-10% of adjusted pMBD (lower 10%, mid 80%, upper 10%) for Korean (all cases, test dataset), reference to mid tertile FS.

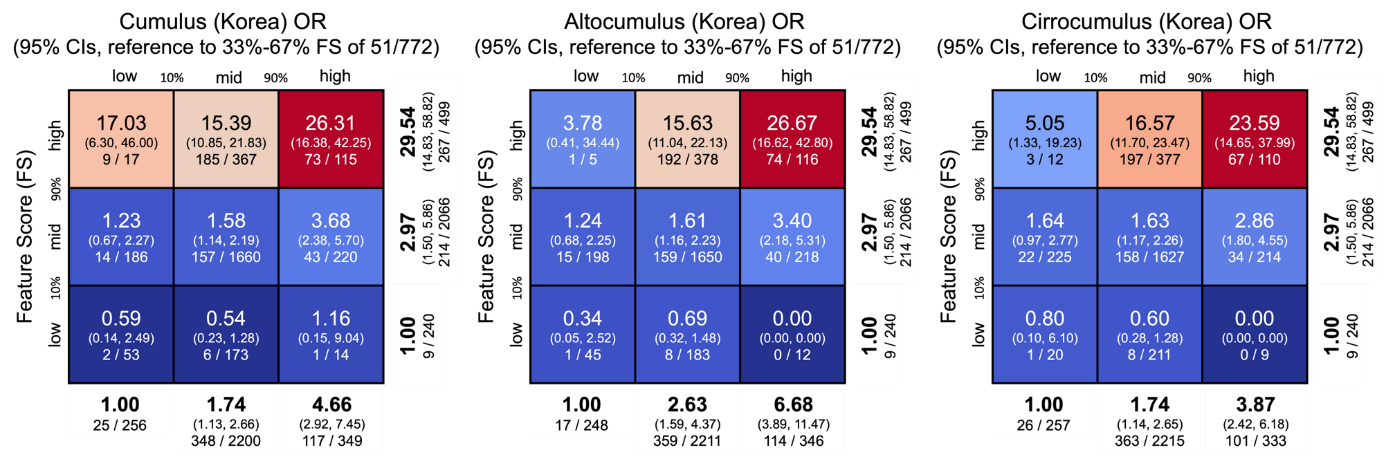

### **Supplementary Fig. 2.**

#### **A.** OR for BC according to tertile of FS and adjusted MBD for Korean (all cases, test dataset) , reference to mid tertile FS.

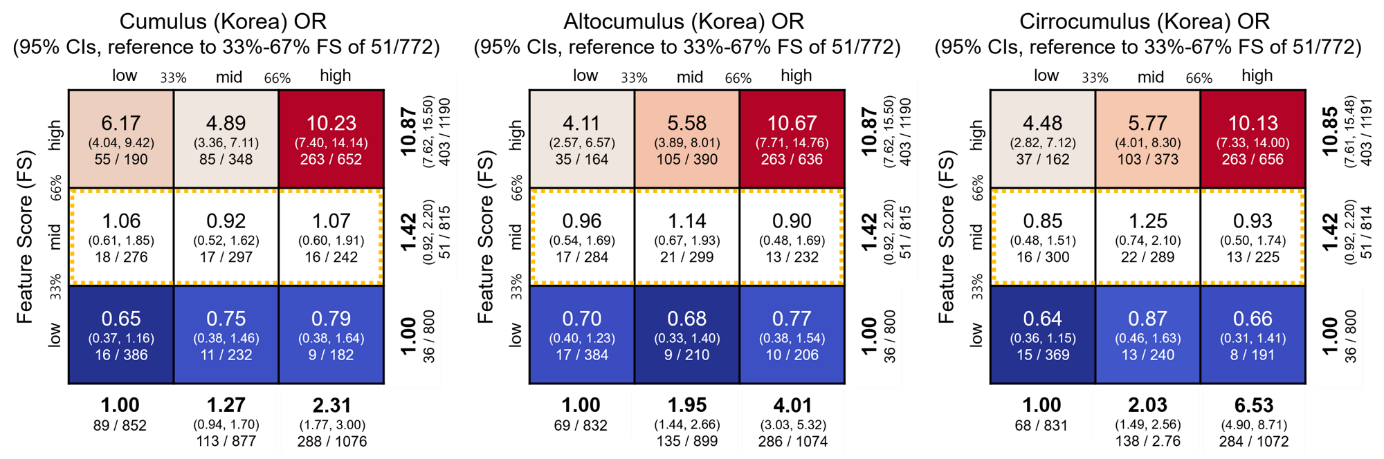

#### **B.** OR for BC according to tertile of FS and adjusted MBD for US White (EMBED, all cases) , reference to mid tertile FS.

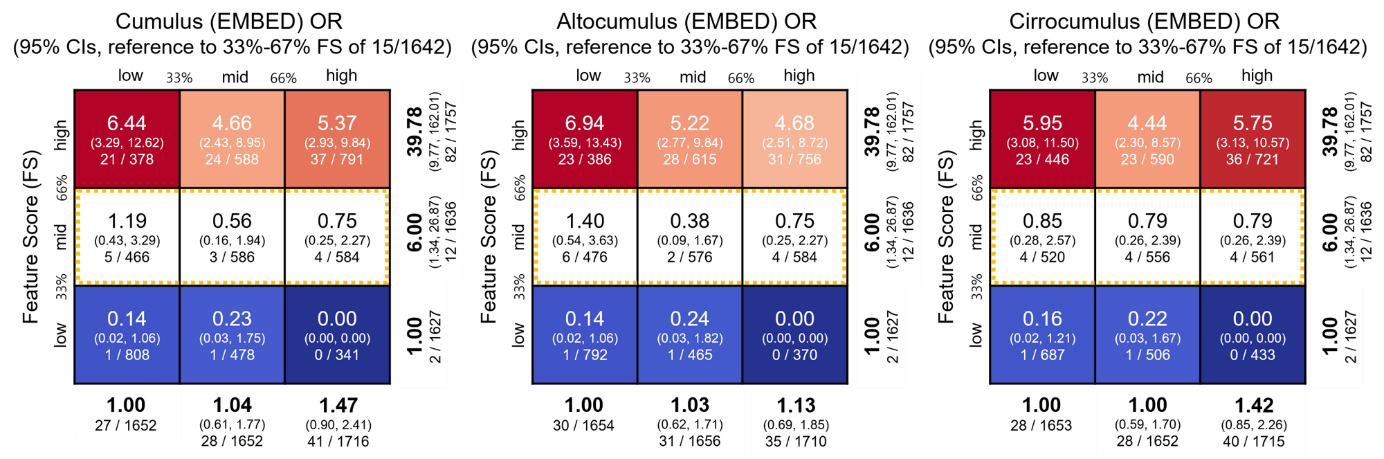

#### **C.** OR for BC according to tertile of FS and adjusted MBD for US White (EMBED, screen-detected cases) , reference to mid tertile FS.

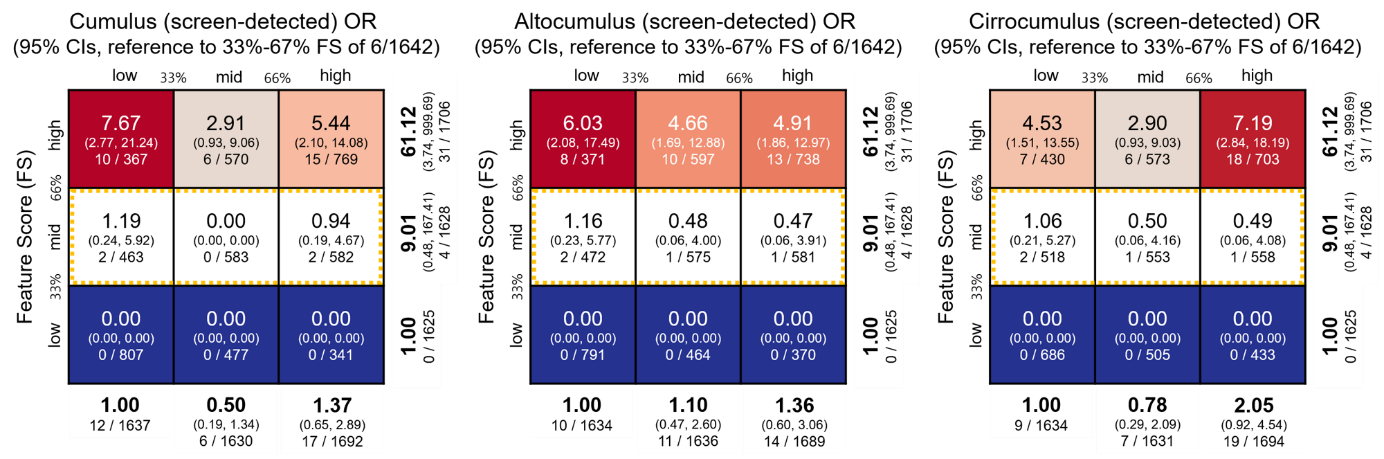

### **D.** OR for BC according to upper-10% of FS (lower 10%, mid 80%, upper 10%) and adjusted MBD tertile for Korean (all cases, test dataset) , reference to mid tertile FS.

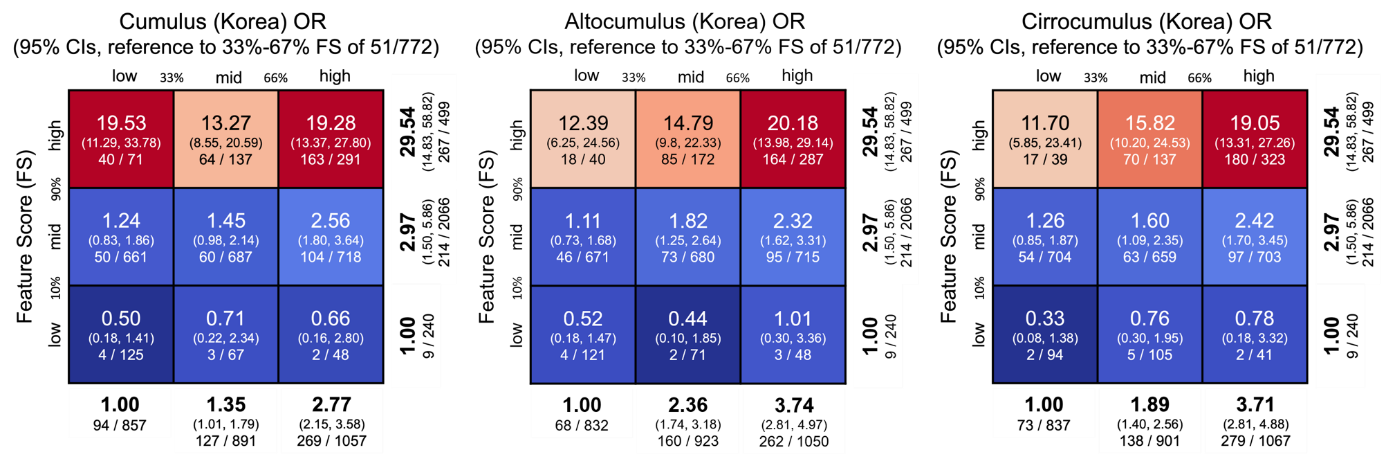

### **E.** OR for BC according to upper-10% of FS (lower 10%, mid 80%, upper 10%) and adjusted MBD tertile for US White (screen-detected cases) , reference to mid tertile FS.

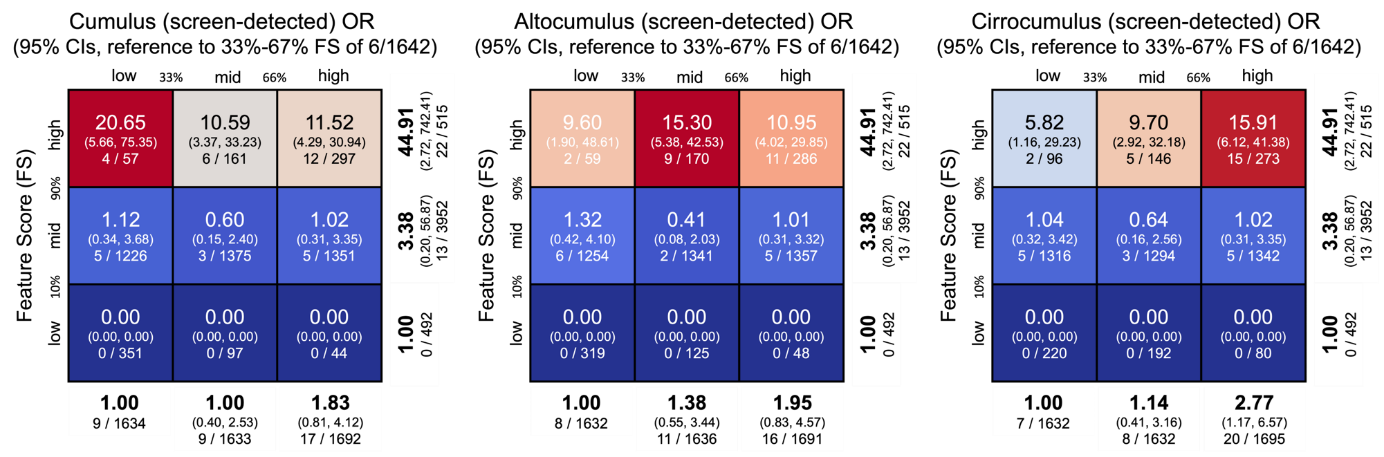

### **F**. OR for BC according to upper-10% of FS (lower 10%, mid 80%, upper 10%) and upper-10% of adjusted MBD (lower 10%, mid 80%, upper 10%) for Korean (all cases, test dataset), reference to mid tertile FS.

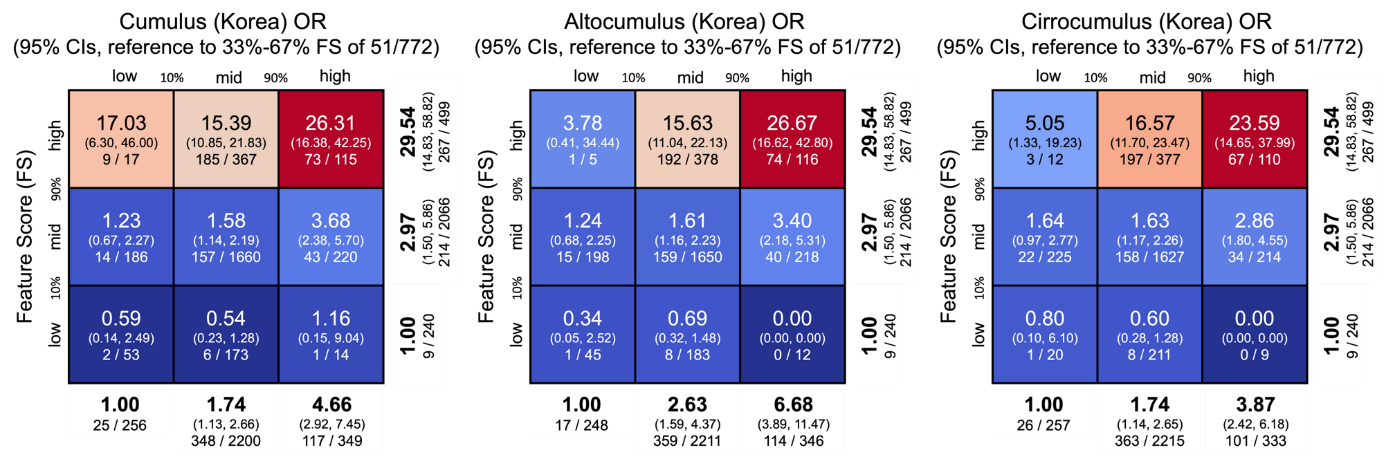
